# Implementing a National Registry of Patients with Inborn Errors of Immunity in Peru: A Mixed-Methods Protocol

**DOI:** 10.1101/2025.11.26.25341085

**Authors:** Liz Veramendi-Espinoza, Kelly De la Cruz-Torralva, Joan Neyra Quijandria, Javier Vargas Herrera, Maritza Puray-Chávez, Percy Soto-Becerra, Leonardo Rojas-Mezarina, Moises A. Huaman, Jacqueline M. Knapke

## Abstract

**Background:** Inborn errors of immunity (IEIs) include more than 500 genetic disorders that cause recurrent infections, autoimmunity, and inflammatory complications. Although classified as rare diseases, their prevalence is increasing, making IEI an emerging public health problem. However, reliable national epidemiological data and coordinated follow-up data of IEI patients are lacking, which potentially affects up to 20,000 people in Peru.

**Objective:** In this study, we aim to describe the protocol of a three-phase mixed-method study that will design, build, and evaluate the usability and feasibility of the first Peruvian National IEI Registry.

**Methods:** Guided by the Exploration-Preparation-Implementation-Sustainment (EPIS) framework, Phase I (qualitative) will map stakeholder needs through in-depth interviews and focus groups; the data will undergo hybrid thematic analysis in ATLAS.ti. Phase II (pre-experimental) will enroll ≥20 clinical immunology consultants and residents to complete standardized tasks on the REDCap-based prototype; efficiency (task time), effectiveness (error rate), and satisfaction will be compared pre/post-iteration. Phase III (observational) will assess feasibility by enrolling ≥126 IEI cases across six national reference hospitals; the primary outcomes are clinical profile, data completeness, and accuracy.

**Results:** The study protocol received institutional review board approval in February 2025 from the UNMSM-Research Ethics Committee. Phase I enrollment started in March 2025. We expect our REDCap-based prototype to incorporate key needs and preferences gathered from focus groups and interviews to develop the platform. By the completion of the study phase II, we will have assessed the usability of the prototype platform. Study phase III will begin in April 2026. The final results are expected by November 2026 and will be published thereafter.

**Conclusions:** By evaluating usability and feasibility in real-world conditions, this protocol lays the groundwork for a national IEI registry. The resulting evidence will inform scale-up decisions and facilitate its incorporation into Peruvian health policy.

**Contributions to the literature:**

- Shows how the EPIS framework can steer real-world adoption of a REDCap-based registry and prospectively measure usability and feasibility across six public hospitals involved in rare-disease surveillance.
- Provides a mixed-methods protocol that designs, iterates and field-tests Peru’s first national registry for inborn errors of immunity, tackling persistent barriers to rare-disease data capture in low- and middle-income countries and offering a scalable blueprint for other resource-constrained settings.
- Outlines a FAIR-aligned governance and data-sharing strategy that can be adapted by future programmes planning registries for rare and orphan diseases in similar contexts.

## Introduction

Inborn errors of immunity (IEIs), formerly “primary immunodeficiencies”, affect an estimated 1:1,200 – 1.2:2,000 individuals worldwide (1,2). Individuals affected by IEI suffer from immune system malfunction and experience recurrent infections, autoimmune diseases, severe allergies, and inflammatory disorders (3). IEI surveillance in Peru remains fragmented: only 287 Peruvian patients (≈3% of the region’s records) have been reported to the transversal Latin American Society for Immunodeficiencies (LASID) registry (4), versus a potential national burden of ≈20,000 cases (5). This, accompanied by the high cost of treatment, places it as an emerging public health problem (6). Timely diagnosis and access to therapeutic interventions such as immunoglobulin replacement or stem-cell transplantation are limited by the absence of structured data (7).

Disease-specific registries close this gap: improve guideline adherence, resource allocation, and research opportunities (8–10). Rare disease registries (RDRs) play a crucial role in advancing medical knowledge and improving clinical care for patients suffering from these conditions. RDRs allow the collection of clinical, genetic, and epidemiological data that would otherwise be unavailable (11). For EII, successful longitudinal models exist in Europe (European Society for Immunodeficiencies - ESID registry) and North America (United States Immunodeficiency Network - USIDNET registry) (12,13). Adapting these experiences to low- and middle-income contexts requires robust evidence on local usability and feasibility.

This project aims to develop an IEI registry in Peru through an implementation sciences framework, which studies the factors, strategies, and approaches that enable the adoption, implementation, and sustainability of evidence-based interventions in health systems (14,15). This framework is embedded within a formative evaluation, an assessment designed to identify contextual barriers and facilitators that guide the adaptation of implementation strategies(16). We expect to bridge the gap between scientific evidence and routine clinical practice, particularly in the Peruvian public health context (14). Implementation strategies will be selected and refined on the basis of formative findings, and later evaluated not only for their effectiveness but also for cost, sustainability, and stakeholder acceptability (15).

The primary objective of this project is to evaluate the usability and feasibility of implementing a registry of patients with Inborn Errors of Immunity. The three secondary objectives/phases are as follows: first, needs assessment, which aims to identify stakeholder requirements for data content, governance, and interoperability. This phase included the design and development of the REDCap-based prototype. The second secondary objective is to conduct a pre-experimental evaluation of the usability of the registry, measuring the efficiency, effectiveness, and user satisfaction of the prototype registry. Finally, the third secondary objective was to evaluate the feasibility of implementing the registry through the outcomes of completeness and describing the clinical-laboratory profile of enrolled Peruvian IEI patients.

## Methods

### Study Overview

The implementation of a nationwide IEI registry requires the contextual depth of qualitative methods and the measurement power of quantitative methods. Accordingly, the project follows a sequential exploratory mixed-methods design (QUAL → QUAN → QUAN) (17), embedded within the EPIS framework (Exploration, Preparation, Implementation, and Sustainability) (14,18), an approach used in implementation science (Figure 1). The study timeline spans February 2025 to November 2026.

**Figure 1.**
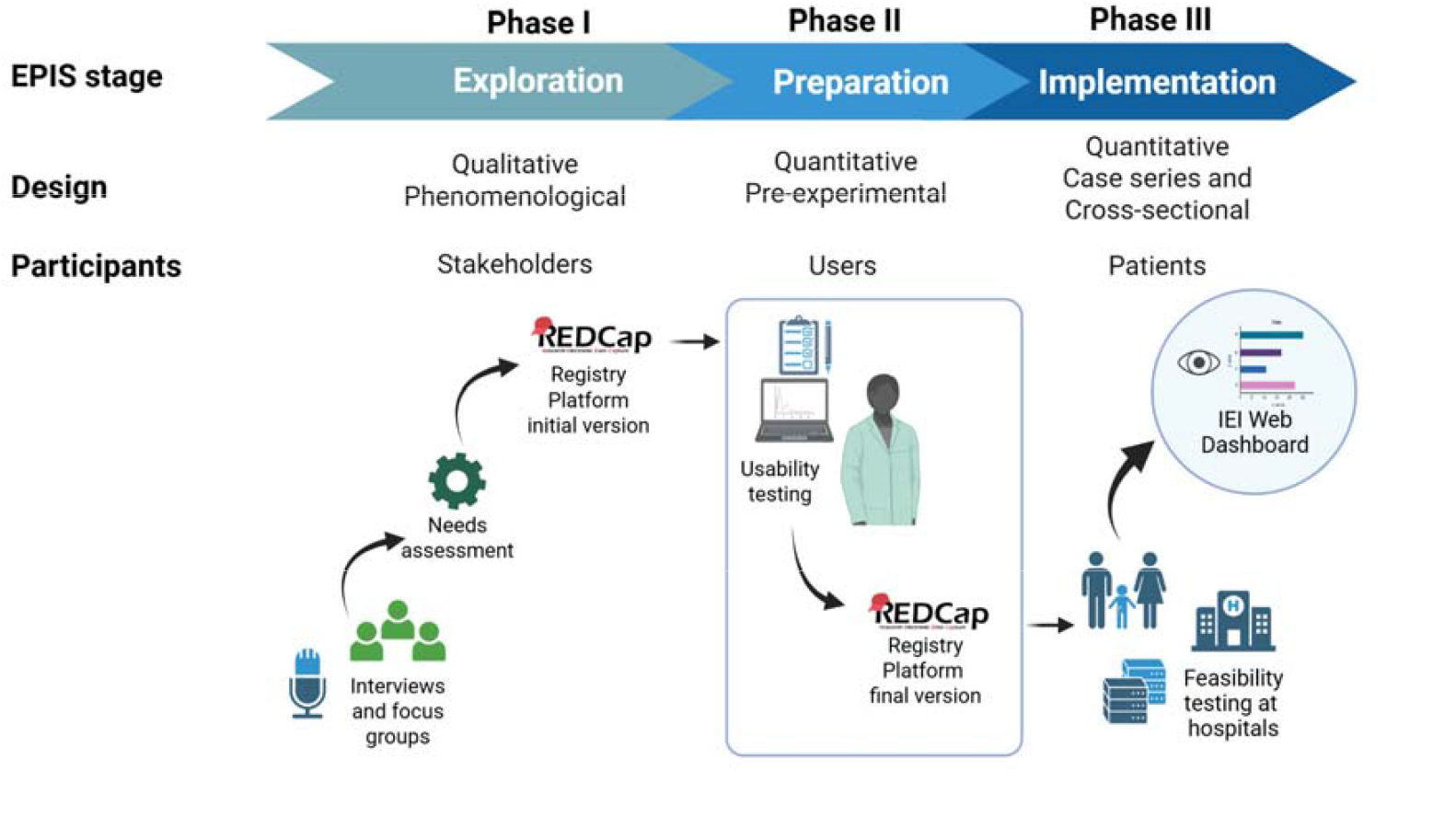
Project design according to the EPIS framework.

In the exploratory phase (Phase I), we will conduct a needs assessment, a tool used to improve resource allocation and design patient-centered health services that align with individual and organizational requirements (19,20). Needs include not only medical aspects, but also social, educational, and financial issues (21). For the preparation phase (phase II), the usability of the designed platform will be assessed. Usability is an essential component for the success of RDRs since a user-friendly system not only improves the user experience but also ensures high-quality data collection and increases patient and professional participation (22,23). For the implementation phase (phase III), monitoring effectiveness outcomes will be assessed through the collection of real patient data.

No previous studies have evaluated these outcomes in existing IEI patient registries; however, there have been previous applications in other chronic diseases such as vitiligo, chronic kidney disease, and mucopolysaccharides (22–24), which provide a solid basis for the evaluation of the implementation of this tool.

### Ethical considerations

The autonomy of the participants will be respected through informed consent. The study will be conducted following the Declaration of Helsinki (25) and the General Health Law (26), respecting the anonymity of the study participants. The data will be handled under Law No. 29733 on the Protection of Personal Data (27), guaranteeing confidentiality and anonymization of the information. This research protocol was approved by the Universidad Nacional Mayor de San Marcos (UNMSM) Research Ethics Committee (Ref FMH-UNMSM/CEI-2025-022).

### Phase I: Needs Assessment for Design and Development of the Registry Overview

A needs assessment is critical for understanding the perceptions of users (doctors, patients, decision makers) based on their individual and collective experiences. Our needs assessment will utilize qualitative methods to identify patterns within qualitative data, allowing us to capture the diversity of perspectives and build useful knowledge for the design of the registry (28).

### Population

We will employ purposive sampling across four stakeholder groups: (I) decision-makers (∼ five participants), (II) national and international IEI experts (∼ eight), (III) clinical users (∼ thirty-four), and (IV) representatives of patient advocacy organizations (∼ five). Recruitment will continue until thematic saturation is reached.

### Data collection and procedures

We will conduct a qualitative observational study combining focus groups and in-depth interviews to assess the needs for implementing the IEI patient registry. Semi-structured interview guides, developed and reviewed by six content experts, will cover feasibility, system structure, navigation, and perceptions of existing registries. Sessions will be audio-recorded with participants’ permission and conducted by a psychologist with expertise in qualitative methods. All recordings will be transcribed verbatim in Spanish using a two-step process: automated speech recognition to generate a draft, followed by dual human verification to correct misrecognitions and remove direct identifiers. Clean, anonymised transcripts will be imported into ATLAS.ti.

### Data analysis

We will use a hybrid (inductive–deductive) thematic analysis. Inductively, two coders will perform open coding on an initial subset of transcripts, compare interpretations, and develop an initial codebook to capture unanticipated patterns and participants’ first-hand perspectives on registry functionalities (29). After iterative coding cycles, we will introduce deductive overlays to enhance explanatory power based on EPIS domains to map context and implementation supports. The resulting hybrid codebook will then be applied to the full corpus. We will generate code reports and analytic matrices to enable constant comparison across stakeholder groups and to identify cross-cutting themes that will inform both the registry’s minimum dataset and preliminary user-interface wireframes.

### Platform infrastructure

The FAIR principles (Findability, Accessibility, Interoperability, Reusability) will guide registry quality attributes (30,31). Development, data capture, and management will be handled with Research Electronic Data Capture (REDCap), a secure, web-based software platform designed to support data collection for research studies(32). REDCap provides an intuitive interface for validated data entry, real-time audit trails, automated export to common statistical packages, and interoperability tools for external data integration. Originally developed at Vanderbilt University, it is now maintained by an international consortium of academic and nonprofit partners(33). The REDCap access will be provided by the Telehealth Unit of the Faculty of Medicine at the UNMSM.

### Reporting standards

Coded segments will be exported to matrix reports for constant comparison and theme development. Reporting will follow the Consolidated Criteria for Reporting Qualitative Research (COREQ) checklist (34).

### Phase II: Pre- and post-Uuability Evaluation Overview

The design will be pre-experimental and quantitative to measure the usability of the system under controlled conditions before its pilot implementation. Usability can be assessed both by qualitative methods, such as focus groups or in-depth interviews(35–37), and by quantitative methods, such as the System Usability Scale (SUS) and Computer Systems Usability Questionnaire (CSUQ) (22,35,36,38). The CSUQ is a quantitative instrument validated in Spanish and previously used in Peru (39,40).

### Population

The study population will be composed of clinical immunologists and clinical immunology fellows in Peruvian hospitals. The total population is 34, distributed in the following hospitals: Guillermo Almenara Irigoyen National Hospital, Edgardo Rebagliati Martins National Hospital, National Institute of Children’s Health - Breña, National Institute of Children’s Health - San Borja, Luis Arias Schreiber Central Military Hospital, and Policia Nacional del Perú “Luis N. Sáenz” National Hospital. There is no single recommendation for sample size in usability studies, as it depends on the context, complexity, and objectives of the study. In general, a group of 5 participants can uncover approximately 80% of usability problems, but this varies depending on the variability in tasks and users (41). For comparative studies or where statistical significance is sought, the recommended optimal range is between 10-12 participants, with a general range of 8-25 (41,42). If a study focuses on complex systems or open tasks, increasing the sample size to ensure greater coverage and complexity of problems is advisable to uncover more serious problems (41,43). Faulkner’s model suggests that 15 users detect ≥90% of usability issues; allowing 15% attrition, we target 20 participants (41). The sample selection will be performed by convenience.

### Data collection and procedure

The participants in this phase will be provided with a username and password to the REDCap platform, as well as a simulated case report, the same for the six hospitals. The time dedicated to the evaluation could be during working hours and in a quiet and adequately equipped environment, which consists of a computer, not necessarily with internet, since REDCap can operate offline. Subsequently, they will be trained in filling out the database. After initial usability testing, identified issues will be prioritised via severity ranking; the prototype will be iterated and retested after prototype optimization. Each user will be their own control.

Usability will be assessed with three complementary measures: task-execution time, number of user errors, and CSUQ version 3 (16 items, reliability of 95%)(44). These metrics capture the efficiency (time), effectiveness (errors), and satisfaction (CSUQ). The CSUQ uses a 7-point Likert scale ranging from 1 “totally satisfied” to 7 “totally dissatisfied”. Consequently, lower scores indicate better perceived usability. The instrument shows a total score (range 16–112) and three subscale scores: system usefulness (Question 1-Q6, score range 6 – 42), information quality (Q7-Q12, range 6 – 42), interface quality (Q13-Q15, range 3 – 21). All four scores (total + three sub-scales) will be analysed separately to provide a granular view of user satisfaction (45,46).

### Data analysis

The choice of tests to analyse and compare these metrics in the initial evaluation (pretest) and final evaluation (posttest) will be made according to the distribution followed by the sample. If it follows a normal distribution, the t-test for paired samples will be used, and if it follows a nonnormal distribution, the Wilcoxon test will be used, which is a non-parametric test for paired data. Analyses will be performed in Stata version 18 with support from the Methodology and Statistics team of the UNMSM Telehealth Unit.

### Phase III: Evaluating Feasibility Overview

Phase III will employ a quantitative observational design to evaluate the feasibility of the IEI registry after its roll-out in the six national referral hospitals that provide Clinical Immunology services in Peru. In registries, feasibility is commonly assessed through the clinical profile, completeness, and accuracy. Together, these indicators will provide an objective measure of how well the registry functions under real-world conditions and whether it is ready for wider scale-up.

### Study population

The study includes all patients with a confirmed IEI diagnosis who have ever attended any of the participating hospitals. From clinic logs, discharge lists, and laboratory or genetic records, the centres collectively identified approximately 205 patients; this cohort constitutes the denominator for all coverage calculations.

### Data-collection procedure

Registry enrollment distinguishes between living and deceased patients. Records for living patients are created only after written informed consent (or parental permission for minors) is obtained. Data from deceased patients are entered under an ethics committee waiver authorized by each institution. Before deployment of the registry, an additional patient list is requested under the Peruvian Transparency Law (47) to establish a baseline against which coverage can later be compared.

During this phase, a comprehensive review of the clinical records of patients with IEI will be carried out at the reference centers selected for the completion of the registry. This review will include information on diagnoses, laboratory tests, treatments, and follow-ups. If essential data are missing, patients or their caregivers will be contacted to complete the record.

### Registry operations and governance

As described in Phase I, the registry will be implemented in REDCap and hosted by the Telehealth Unit of the Faculty of Medicine at UNMSM, which already maintains an institutional REDCap instance on a secure, university-managed server. A dedicated data manager from the unit will configure the project, implement role-based permissions, perform quarterly backups, and store on a geographically separate UNMSM server to guarantee data availability in the event of local failure. Trained clinicians or residents will enter patient information directly from source charts through the web interface; they will receive a concise, standardized data-entry manual and access to a help-desk e-mail. User privileges follow a “minimum-necessary” model: clinicians possess data-entry and edit rights limited to their own hospital, whereas the central data manager alone retains design and export permissions. Finally, automated integrity scripts run each week to flag impossible dates, out-of-range laboratory values, and orphan records; the resulting queries are returned to site staff for resolution. Together, these measures ensure that registry operations comply with Peru’s Personal Data Protection Law (Law 29733) while maintaining the confidentiality, integrity, and availability of patient data throughout the study lifecycle.

### Data analysis

Regarding the clinical profile, descriptive statistics (means, medians, and proportions) will summarise the data. Furthermore, three data-quality outcomes will be assessed. Coverage is defined as the proportion of the 205 eligible patients whose core record is created in REDCap. Completeness refers to the percentage of mandatory data elements completed in each record and will be summarised as the mean completeness with a 95% confidence interval. Accuracy represents the proportion of audited data elements that match the information in the source chart and will be evaluated in a random subsample of records.

For the completeness and accuracy audit, the required sample size was calculated with OpenEpi (version 3.01). Assuming an expected completeness of 70%(48), a precision of ±5%, and a confidence level of 95%, and applying a finite-population correction for N = 205, we determined that 126 patient records are needed. Feasibility will be deemed acceptable if field completeness reaches at least 70%. Sensitivity analyses will explore variability across hospitals. Updated records extracted from the registry—diagnoses, laboratory tests, and treatments—will feed a web-based epidemiological-surveillance dashboard. A flow diagram of the data life- cycle—from patient identification and consent through data entry, quality control, export, and dashboard display—will be finalised during Phase I (registry design) and included as a figure in the results section.

## Results

The proposed work was awarded funding support in March 2024 through CONCYTEC and PROCIENCIA, the national funding agencies for research in Peru. The project received institutional review board approval in February 2025 from the UNMSM-Research Ethics Committee (FMH-UNMSM/CEI-2025-022). Phase I enrollment started in March 2025. We will incorporate stakeholder requirements gathered from focus groups and interviews into the REDCap-based prototype. Phase II is scheduled from December 2025 to March 2026, and we will have assessed the usability of the prototype platform. Study phase III will begin in April 2026. The final results are expected by November 2026 and will be published thereafter.

## Discussion

Inborn errors of immunity are clinically significant conditions that demand timely diagnosis, coordinated follow-up, and equitable access to specialized care. Currently, a harmonized, national-level data source that can inform clinical decision-making, resource allocation, and research priorities for this population is lacking. Therefore, establishing an electronic registry that adheres to international data-quality standards represents a crucial step toward improving patient outcomes, fostering local research capacity, and aligning the country with global IEI surveillance efforts.

Several factors indicate that the project is achievable within the Peruvian health-care context. The use of REDCap—a secure, widely adopted, and regulation-compliant platform—simplifies data capture and management while ensuring interoperability with future research initiatives. Its implementation in six referral hospitals allows the team to measure usability, identify technical or workflow barriers, and adjust the system before national roll-out. Finally, evaluating usability with well-established indicators of efficiency, effectiveness, and satisfaction, and employing the Spanish-validated CSUQ provides a rigorous framework for assessing user experience.

The study’s success depends on active engagement from clinicians, decision-makers, and patients. Limited availability or competing demands on participants’ time may restrict data entry and reduce the representativeness of needs assessment and usability findings. Securing institutional permissions for chart access is essential, but may be delayed by administrative processes. Compliance with Peru’s Personal Data Protection Law (27) imposes strict requirements on data handling, and heterogeneous record-keeping systems across hospitals could compromise the completeness and timeliness of extracted information. In addition, obtaining fully informed consent may constrain the number of patients included in the registry, thereby affecting sample size and the breadth of clinical data captured.

Registry studies typically measure effectiveness outcomes through various parameters, such as the number of patients enrolled, the availability of information, the development of research projects, enrollment of patients in clinical trials, and clinical information (49,50). However, these outcomes will be evaluated after the initial implementation since they need to be longitudinal. In the long term, the implementation of registries requires improved capacity to collect high-quality information. The path is toward the automation of data extraction from hospital information systems to facilitate the collection of data from all patients and reduce the need for manual processing, which is costly in terms of human resources and increases the possibility of human error (10).

## Data Availability

All data produced in the present study are available upon reasonable request to the authors

## Declarations

### Ethics approval and consent to participate

The study protocol (Ref FMH-UNMSM/CEI-2025-022) was reviewed and approved by the Research Ethics Committee of the Faculty of Medicine, Universidad Nacional Mayor de San Marcos (UNMSM). All Phase I and Phase II participants will provide written informed consent prior to data collection. For Phase III, written informed consent (or parental permission for minors) will be obtained before any living patient’s data are entered in the registry; the committees waived individual consent for deceased cases because only retrospective, de-identified chart data are captured. The project adheres to the Declaration of Helsinki and to Peruvian Law No 29733 on Personal-Data Protection.

### Consent for publication

No individual person-level images, videos or identifiable details are presented in this manuscript; therefore, consent for publication is not applicable.

### Availability of data and materials

Because the registry will contain clinically sensitive information on a rare-disease population, the full dataset cannot be placed in a public repository. De-identified aggregated tables and the REDCap data dictionary will be available in Zenodo. Investigators may request the underlying anonymised dataset from the corresponding author (LVE) via a data-sharing agreement.

## Authors’ contributions

LVE conceived the study, drafted the protocol and is principal investigator. KDC and LRM advise the REDCap architecture and will oversee data quality. JVH and JNQ provide clinical oversight and methodological guidance. PSB and MPC contribute to statistical design and will analyse quantitative data. MAH and JMK advise on qualitative methods and mixed-methods integration. All authors reviewed the manuscript critically for important intellectual content, approved the final version, and agreed to be accountable for all aspects of the work.

## Acknowledgements

This work is funded by the Consejo Nacional de Ciencia, Tecnología e Innovación Tecnológica (CONCYTEC) and the Programa Nacional de Investigación Científica y Estudios Avanzados (PROCIENCIA) within the framework of the E077-2023-01-BM “Becas en Programas de Doctorado en Alianzas Interinstitucionales” contest, grant number (PE501090535-2024) and the E033-2023-01-BM “Alianzas Interinstitucionales para Programas de Doctorado” contest, grant number (PE501090535-2024).

We also thank Dr Abisag Durand Guevara and Dr Luis Gamero Oviedo, who lead the Peruvian Registry for Rare and Orphan Diseases, for their support and openness to collaborate with this registry project.

## Authors’ information

LVE is a doctoral candidate in Health Sciences at the Universidad Nacional Mayor de San Marcos (UNMSM); the present study constitutes her doctoral thesis project.

